# Comparative fine-mapping of breast cancer susceptibility loci using summary statistics methods and multinomial regression

**DOI:** 10.64898/2026.04.21.26351364

**Authors:** Denise G. O’Mahony, Jonathan Beesley, Maria Zanti, Joe Dennis, Diptavo Dutta, Peter Kraft, Vessela Kristensen, Georgia Chenevix-Trench, Douglas F. Easton, Kyriaki Michailidou

## Abstract

Summary statistics fine-mapping methods offer advantages over classical methods, including avoiding data-sharing constraints and improved modelling of correlated variables and sparse effects. However, its performance has not been comprehensively evaluated in breast cancer using real-world data. Previous multinomial stepwise regression (MNR) fine-mapping analyses for breast cancer identified 196 credible sets. Here, we apply summary statistics fine-mapping, compare methods, and assess parameters influencing performance. Using summary statistics from the Breast Cancer Association Consortium, we compared *finiMOM, SuSiE*, and *FINEMAP* to published MNR results across 129 regions. Performance was assessed by recall using in-sample and out-of-sample LD. Discordant credible sets were examined for technical factors, and target genes were defined using the INQUISIT pipeline. *SuSiE* showed the closest agreement with MNR. Results varied across regions depending on the assumed number of causal variants (*L*), with higher values reducing recall and no single *L* maximising performance. At optimal *L* per region, *SuSiE* identified 8,192 CCVs in 244 credible sets, with recall of 88%, 86%, and 72% for overall, ER-positive, and ER-negative breast cancer. Thirty MNR sets were missed. Discordance was partially explained by allele flips, imputation quality, and array heterogeneity. Fifty-two MNR-identified genes, including *BRCA2, WNT7B* and *CREBBP* were not recovered, while additional candidate genes were identified. Using out-of-sample LD reduced recall by 3% but identified novel variants. Fine-mapping results vary across methods, and no single approach is sufficient. The choice of *L* strongly influences results, and combining analytical approaches with functional validation can improve causal variant identification.

## Introduction

Genome-wide association studies (GWAS) have identified many risk-associated single nucleotide polymorphisms (SNPs) that contribute to cancer susceptibility. In breast cancer, collaborative efforts of the Breast Cancer Association Consortium (BCAC) and the development of breast cancer-specific genotyping arrays, such as the iCOGS and OncoArray, have led to the discovery of >210 susceptibility loci that account for approximately one fifth of the familial relative risk of the disease^[1,2]^.

Although GWAS have been successful at detecting risk loci, identifying the underlying causal variants within risk regions is complicated by the strong correlations among variants due to linkage disequilibrium (LD). In addition, multiple independent variants within the same region may be disease associated. Fine-mapping analyses aim to identify likely causal variants by grouping correlated variants into independent “credible sets”, each of which is expected to contain one or more candidate causal variants (CCVs). Distinguishing likely causal variants within a credible set typically requires orthogonal in-silico or in-vitro data to assess functional effects.

A classical approach to defining credible sets uses stepwise regression to estimate the number of independent risk variants, followed by likelihood comparisons (where each SNP is compared to the most strongly associated variant), while conditioning on lead variants in other sets to determine the variants that cannot be excluded from the credible set. In breast cancer, the largest fine-mapping analysis to date was conducted by dense genotyping and imputation of 150 breast cancer-associated regions, followed by multinomial stepwise logistic regression (MNR) across ER-neutral, ER-positive and ER-negative disease in >217,000 subjects of European ancestry from BCAC^[3]^. This analysis identified 196 strong evidence credible sets in 129 regions, with in-silico predictions implicating 191 likely target genes.

The stepwise approach can identify multiple causal variants and MNR enables joint modelling of multiple subtypes or outcomes (e.g., ER-positive versus ER-negative breast cancer) within a unified framework. However, numerous Bayesian fine-mapping methods have recently emerged that offer several potential advantages over frequentist methods, including posterior inclusion probabilities (PIPs), probabilistic credible sets (containing the true causal variant(s)) enabling direct comparison of mapping precision across loci, improved credible set refinement in regions of high linkage disequilibrium (LD) and the ability to incorporate prior biological information^[4–6]^. Many Bayesian methods also rely on GWAS summary statistics, increasing their applicability across traits when individual-level data are unavailable^[7,8]^. Due to the lack of ground truth, these methods are often evaluated on simulated data^[7,9–12]^, which while convenient, it may not capture the complexity of real-world genetic architecture, particularly for heterogeneous traits such as cancer. Moreover, applications to empirical data commonly prioritise performance such as fine-mapping accuracy and calibration over biological interpretation and often focusing on traits with relatively simpler genetic architectures^[10,13–15]^.

In this work, we applied Bayesian summary statistics fine-mapping pipelines to 129 previously identified breast cancer risk regions^[3]^ and compared them to MNR, which represents the most comprehensive published fine-mapping analysis in breast cancer and defines the prevailing causal variant sets for the disease. We compared methods in terms of causal variant prioritisation, methodological differences and statistical uncertainty using posterior inclusion probabilities. Analyses were based on summary statistics data from the largest available breast cancer association study in women of European ancestry, based on >217,000 cases and controls. The impact of in-sample versus out-of-sample LD matrices was also assessed.

## Materials and Methods

### Summary statistics

GWAS summary statistics generated by Fachal *et al*. 2020^[3]^ were used as input to fine-mapping. Briefly, summary statistics data were based on individuals of European ancestry from studies participating in BCAC. Samples were genotyped using the OncoArray^[16]^ and iCOGS^[17]^ custom arrays and imputed against the 1000G Project reference panel (Phase 3). The large majority of cases and controls were drawn from population-based case-control studies or nested case-control studies within population-based cohorts, with a small subset drawn from clinic-based studies with ascertainment based on family history of breast cancer. Summary statistics (per-allele odds ratios and standard errors) were generated separately from iCOGS and OncoArray data and combined by fixed-effects meta-analysis in METAL. Summary statistics for three phenotypes were analysed: (1) Overall breast cancer (88,937 controls and 109,900 cases), (2) Estrogen receptor (ER)-positive breast cancer (88,230 controls and 67,136 cases) and (3) ER-negative breast cancer (88,572 controls, 17,506 cases). For each phenotype, variants were filtered according to minor allele frequency (MAF) and statistical significance. Common variants (MAF ≥ 0.01) were retained when P ≤ 0.05, whereas low-frequency variants (0.001 ≤ MAF < 0.01) were retained when P ≤ 1×10^−4^. Multi-allelic variants and variants with imputation quality score < 0.3 were excluded from all analyses.

### Linkage disequilibrium matrices

LD matrices from two datasets were used. First, partially in-sample LD dosage matrices computed from individual-level data from the OncoArray dataset, generated using the LDstore2 software^[18]^. Since the summary statistics were derived from a meta-analysis of OncoArray and iCOGS datasets, this LD represents partial in-sample LD. Second out-of-sample LD matrices based on individuals of British-ancestry from the UK Biobank, were retrieved from https://broad-alkesgroup-ukbb-ld.s3.amazonaws.com/.

### Fine-mapping pipelines

We compared the fine-mapping approaches *FINEMAP*^[19]^, *FiniMOM*^[11]^ and *SuSiE*^[7,20]^ to the published MNR fine-mapping results, generated based on the same data (Fachal *et al*. (2020)^[3]^). Both in-sample and out-of-sample LD matrices were used in this assessment. The pipelines were applied to 129 of the 150 genomic regions previously fine-mapped by MNR^[3]^ and fine-mapping was performed for overall breast cancer, ER-positive subtype and ER-negative subtype. Regions were selected based on the presence of high confidence causal sets (containing at least one associated variant at P < 1×10^−6^ and referred as ‘signals’ in Fachal *et al*., 2020), while regions containing only moderate-confidence causal sets (n=21) were excluded. Regions were defined by taking 1 Mb window around the identified “index” variants (i.e., most statistically significant variant in each region).

*FINEMAP* was applied with the default settings while using the shotgun stochastic search (SSS) algorithm and allowing for exactly 10 causal variants per region. *finiMOM* was applied also allowing a maximum of 10 causal variants per region, with automatic local approximation (ALA), default model size prior hyperparameter (u = 2) and a purity threshold of 0.5. *SuSiE* (Sum of Single Effects) was applied using the regression summary statistics (RSS) algorithm, allowing up to 10 causal variant effects (L = 10), at 1000 iterations and estimating the prior and residual variances. In all pipelines the following input data were used: standardised summary statistics (z-scores), LD matrices and sample sizes, while SNP-wise posterior inclusion probabilities (PIPs) were produced at a 95% credible set threshold.

### Comparison of Bayesian fine-mapping methods against the multinomial model

The Bayesian fine-mapping methods *FINEMAP, SuSiE* and *finiMOM* using in-sample LDs, were compared against published fine-mapping results obtained using MNR in Fachal *et al*. 2020^[3]^. Comparison analyses were conducted per pipeline using summary statistics for overall breast cancer, ER-positive and ER-negative disease. For consistency, results from the overall breast cancer analysis were compared with ER-neutral MNR results (defined in Fachal *et al*. 2020 as regions with no significant difference between the ER-positive and ER-negative subtypes), while ER-positive and ER-negative analyses were compared to subtype-specific MNR results. Overlap in CCVs and credible sets was determined across methods. The main comparison metrics measured between the Bayesian methods and MNR was CCV and credible set recall. CCV recall was defined as the number of Bayesian-derived CCVs overlapping MNR-derived CCVs, divided by the total number of MNR CCVs for each phenotype (5,510 for ER-neutral, 1,238 for ER-positive and 646 for ER-negative subtype). Credible set recall was defined as the number of overlapping credible sets between each Bayesian summary statistics method and MNR (containing at least one CCV of the MNR credible sets), divided by the total number of MNR credible sets also called ‘signals’ in Fachal *et al*. (n=196; 101 for ER-neutral, 66 for ER-positive and 29 for ER-negative).

*SuSiE* was selected for further evaluation on the basis of accuracy, interpretability and recall to the MNR results^[3]^. Further comparison of *SuSiE* against MNR results was performed by running the model with the L parameter fixed at values between 1 to 50, where L represents the maximum number of causal variants assumed per region. At these L values, CCV and credible set recall was evaluated as before. The proportion of regions working was also defined, corresponding to the number of regions containing any credible sets, irrespective of overlapping credible sets, divided by the total number of regions for each phenotype based on Fachal *et al*. 2020 (86 for ER-neutral, 47 for ER-positive and 23 for ER-negative subtype). To account for heterogeneity in results across different L, for each region we selected fine-mapping results corresponding to the value of L that maximised recall with MNR. Based on these results, CCV recall was stratified by region complexity; regions with one MNR credible set, regions with three MNR credible sets and all 129 regions analysed. In addition to recall, we also measured precision across L parameters and phenotypes, defined as the number of overlapping credible sets or CCVs between *SuSiE* and MNR divided by the total number of sets or CCVs defined by *SuSiE*. The Jaccard index was also computed, defined as the number of overlapping credible sets or CCVs by the total number of unique CCVs or sets across both *SuSiE* and MNR. To examine how our *SuSiE*-inferred posterior inclusion probabilities (PIPs) relate to previously reported fine-mapping results, we performed a variant-level comparison between *SuSiE*-derived credible sets and MNR-derived CCVs. Specifically, variants identified by *SuSiE* were grouped into PIP bins (0-0.1, 0.1-0.2, …, 0.9-1.0) and for each bin we calculated the proportion of variants that overlapped with MNR-derived CCVs. The median size of the credible sets generated by each method was also compared. Benchmarking was also performed separately using in-sample and out-of-sample LD matrices from the UK Biobank. Differences in credible set size between in-sample and out-of-sample LD matrices were assessed separately for each phenotype using two-sided Wilcoxon rank-sum tests, with statistical significance evaluated at a threshold of P<0.05. Potential allele flips between observed summary statistics versus expected z-scores from LD matrices were explored using *SuSiE’s kriging*_*rss* algorithm. To assess array-specific heterogeneity, a sensitivity fine-mapping analysis was performed using summary statistics from iCOGS and OncoArray separately. Distributional differences were assessed using two-sided Wilcoxon rank-sum tests, with Benjamini-Hochberg-adjusted P-values (P<0.05).

### Detection of miscalibrated loci with SLALOM

To assess the reliability of our meta-analysis fine-mapping results (based on the OncoArray and iCOGS), we annotated the breast cancer risk regions selected for fine-mapping using previously published SLALOM results from Kanai et al. (2022)^[21]^. SLALOM identifies regions with miscalibrated posterior inclusion probabilities arising from violations of modeling assumptions, such as allelic heterogeneity, misspecified LD or heterogeneous effect sizes, by assessing whether the observed pattern of association statistics is consistent with a single causal variant model. Regions flagged as suspicious by SLALOM in the original study were considered potentially unreliable.

### Gene target prediction

Gene targets for identified CCVs were predicted using the breast cancer gene prediction pipeline, INQUISIT^[1,3]^, which integrates multiple layers of genomic data through a scoring framework for distal variants, proximal regulatory variants and coding variants. CCV positions were intersected with chromatin interaction data from capture Hi-C experiments across six breast cell lines, as well as ChIA-PET and Hi-C analyses. Enhancer-promoter interactions were evaluated using datasets from PreSTIGE^[22]^, IM-PET^[23]^, FANTOM5^[24]^ and super-enhancers^[25]^. Additionally, breast tissue eQTL data from European ancestry samples were utilized for variants within 129 genomic intervals using TCGA^[26]^, METABRIC^[27]^ and NHS datasets^[28]^. Transcription factor and histone modification ChIP-Seq data from ENCODE^[29]^ and Roadmap Epigenomics^[30]^ were also considered, alongside genomic features identified as significantly enriched. Furthermore, gene expression RNA-Seq data from various breast cancer lines and normal samples were used, as well as topologically associated domain boundaries from T-47D cells. To assess the impact of intragenic variants, the Variant Effect Predictor (VEP)^[31]^ of Ensembl was implemented, incorproating predictions from SpliceAI^[32]^, MaxEntScan^[33]^ and dbscSNV^[34]^ for splicing variants and PolyPhen^[35]^, SIFT^[36]^, REVEL^[37]^ and dbNSFP^[38]^ for missense variants. Variant predictions affecting post-translational modifications were derived from the PTMsnp database^[39]^. The gene recall rate was calculated similarly to before, by dividing the number of INQUISIT level 1 genes derived from *SuSiE*-annotated CCVs by the INQUISIT level 1 genes annotated to MNR-derived CCVs.

### Systematic review of experimentally validated fine-mapped variants

To systematically evaluate fine-mapping results against experimentally supported causal variants, we compiled functionally validated breast cancer variants from over 25 breast cancer published fine-mapping studies. Variants were mapped to exact genomic coordinates and cross-referenced only if there was direct experimental evidence supporting their functional impact, including luciferase reporter assays, eQTL analyses (TCGA, METABRIC), transcription factor binding or chromatin interaction studies. Two variant sets were specifically analysed: (i) new CCVs detected for the first time in *SuSiE* credible sets not found by MNR and (ii) MNR CCVs that were missed by *SuSiE*.

### In-silico functional annotation

For each *SuSiE*-derived credible set, CCVs were assessed on their putative functional significance by applying a heuristic score using RegulomeDB^[40]^. Publically available DNase hypersensitivity, transcription factor binding and histone modification ChIP-seq data from the ENCODE project^[29]^ and *ReMap*^[30]^ were also used to overlay functional annotations on each set.

## Results

### Comparing Bayesian fine-mapping pipelines to MNR

We compared results from the Bayesian fine-mapping pipelines *FINEMAP, FiniMOM* and *SuSiE*, applied to meta-analysis summary statistics from iCOGS and OncoArray with in-sample LD matrices derived from the OncoArray, to previously published fine-mapping results by Fachal *et al*., (2020) using multinomial regression (MNR) on the same dataset^[3]^. An overview diagram of the study is provided in **Fig.1**. CCV overlaps between pipelines, including multinomial regression results, are presented in **Fig. 2A**. Overall, the methods yielded distinct fine-mapping results, with *FINEMAP* identifying 82,998 CCVs, *SuSiE* 9,601 CCVs and *finiMOM* 3,515 CCVs. Most causal variants identified by *SuSiE, finiMOM* and MNR were shared with other pipelines, whereas the majority of CCVs by *FINEMAP* were unique. Only 294 CCVs were overlapping across all four pipelines, while 5,379 were common between MNR, *SuSiE* and *FINEMAP*. **Fig. 2B** summarises the recall of MNR-derived credible sets by the Bayesian methods per phenotype, indicating variable alignment between methods. *FINEMAP* yielded the highest credible set recall rate (96% across all phenotypes), followed by *SuSiE* (81%) and *finiMOM* (79%). Based on these results and the substantial inflation in CCVs produced by *FINEMAP, SuSiE* was selected for further assessment in comparison to MNR.

**Fig. 1.**
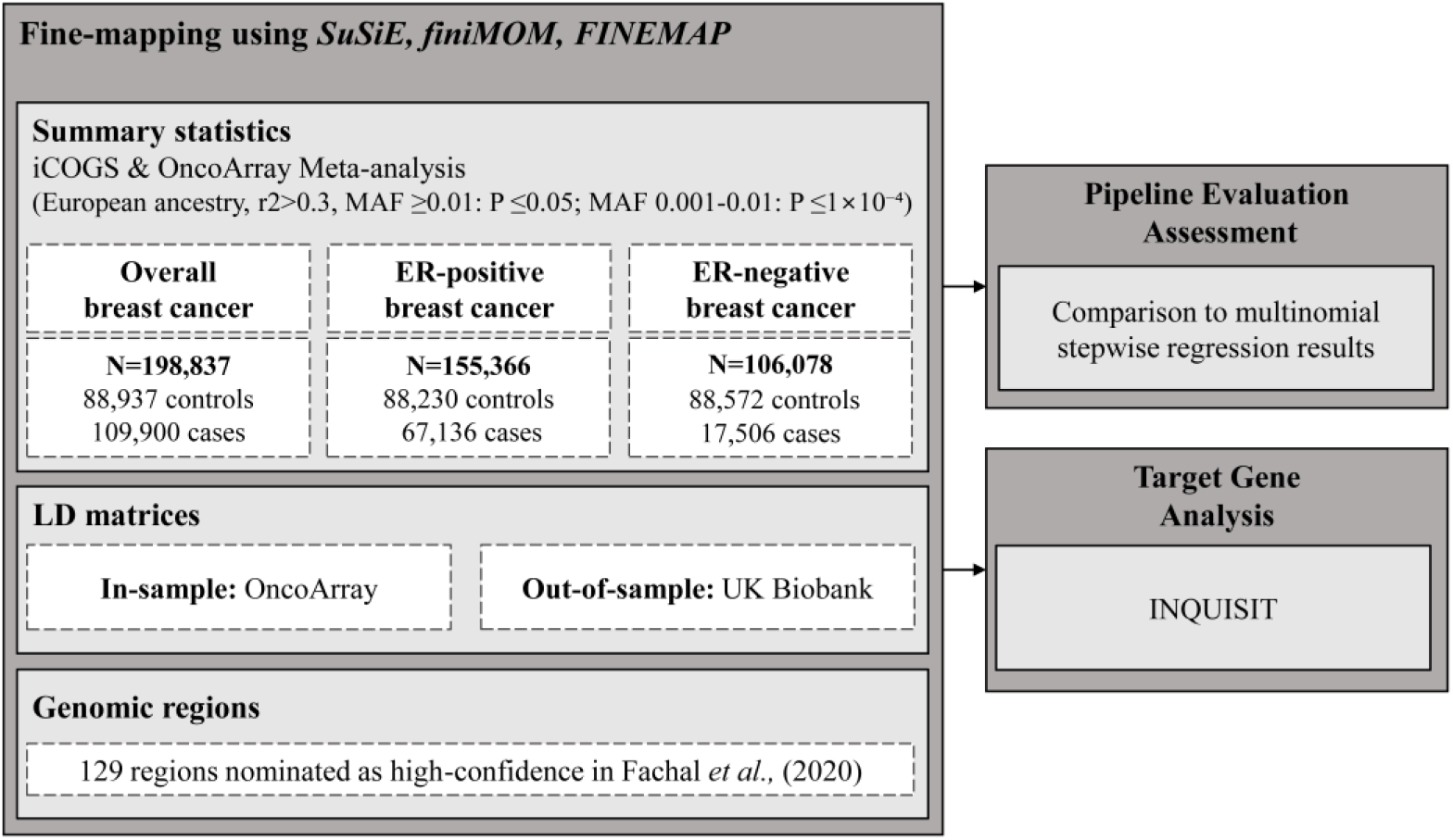
Flowchart of study design. Fine-mapping was performed using three Bayesian summary statistics fine-mapping pipelines: *SuSiE, FINEMAP* and *finiMOM*. Summary statistics data for Overall breast cancer, ER-positive and ER-negative subtypes, based on European ancestry were used in all analyses. Both in-sample LD matrices and out-of-sample LD datasets were incorporated for comparison. Fine-mapping was conducted in association to the three subtypes, across 129 genomic regions nominated by Fachal *et al*., (2020) at high-cofidence. Identified credible sets and CCVs were used to compare the three Bayesian approaches against known candidates identified by multinomial stepwise regression (Fachal *et al*. 2020). Nominated CCVs we analysed further on potential targets. MAF, minor allele frequency; ER, estrogen receptor; N, number; LD, linkage disequilibrium; CCV, candidate causal variants.

**Fig. 2.**
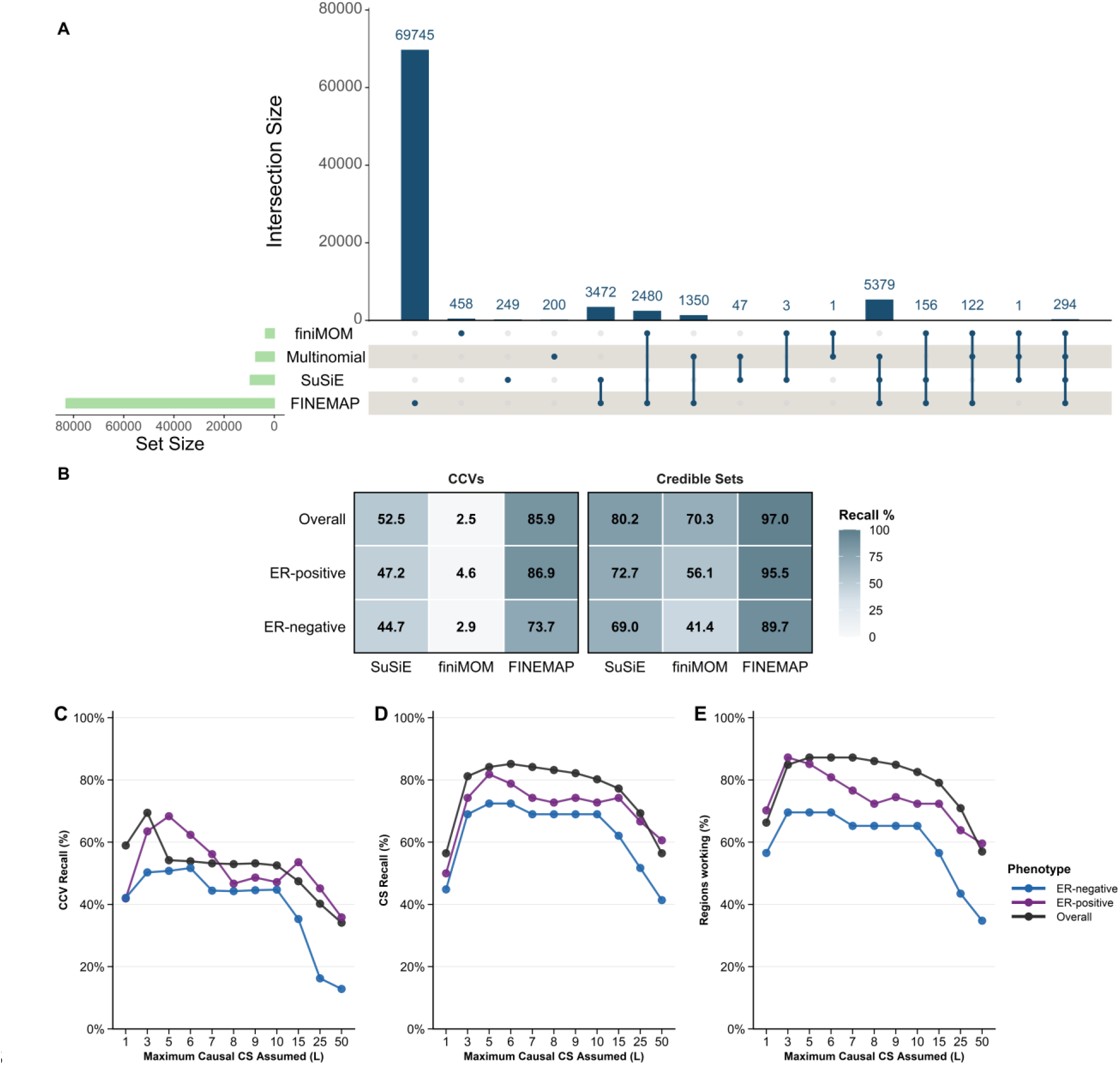
CCV overlap between *finiMOM, SuSiE* and *FINEMAP* and comparison against MNR. (**A**) Overlap of CCVs between *finiMOM, SuSiE, FINEMAP* and published results using multinomial regression across phenotypes. The UpSet plot displays the total number of CCVs identified by each fine-mapping pipeline (green bars), with unique CCVs shown by single dots and shared CCVs represented by interconnected dots. The counts of unique or shared entries are shown by the blue bars. (**B**) Recall of MNR-derived CCVs and credible sets by the three Bayesian methods, per phenotype. (**C**) Recall of MNR-derived CCVs detected by *SuSiE*, coloured by ER subtype, across L. (**D**) Recall of MNR-derived credible sets detected by *SuSiE*, coloured by ER subtype, across L. (**E**) Percentage of regions with MNR-derived credible sets by *SuSiE*, coloured by subtype, across L. CCV, candidate causal variants; CS, credible sets; MNR, multinomial regression.

### *SuSiE* vs Multinomial Regression: Detailed Assessment

*SuSiE’s* robustness and alignment to MNR was assessed by fixing the L parameter to variable number of causal variants per region (between 1-50). **Fig. 2** summarizes CCV recall (**Fig 2C**), independent credible set recall (**Fig. 2D**) and region-level recall by *SuSiE* (**Fig. 2E**) across L (i.e., maximum number of credible sets assumed per region). CCV recall ranged from 18-69%, indicating moderate recall of CCVs. Credible set recall was higher, reaching 83% for overall breast cancer at L = 6, 81% for ER-positive at L = 5 and 74% for ER-negative at L=5. The percentage of regions identifying at least one credible set reached 88% (76/129) for overall breast, 88% for ER-positive (41/47) and 70% for ER-negative breast cancer (16/23). Importantly, the number of regions yielding any fine-mapping results did not increase monotonically with the number of assumed causal variants (L): some regions produced credible sets only at specific L values, with some losing credible sets at higher L, while others only began to yield results at larger L. Overall, CCVs and credible set recall as well as regions working declined with the number of causal variants assumed across loci. Given this heterogeneity in results, for each region, we selected the fine-mapping results that achieved the highest MNR credible set recall across L (**Supplementary Table S1 & S2**).

Using these highest-recall results, CCV and credible set recall for each phenotype were stratified by complexity, defined by the number of independent credible sets identified by MNR in Fachal et al., (2020): (i) for all regions, (ii) regions containing a single MNR credible set and (iii) regions consisting of three MNR credible sets (**Fig. 3A**). In regions with single MNR sets, recall by *SuSiE* was 83% for overall breast cancer, 72% for ER-positive and 77% for ER-negative. In the same regions, *SuSiE* identified multiple independent sets in a subset of cases, redefining 12/53 (23%) of overall disease regions, 5/25 (20%) of ER-positive and 3/13 (23%) of ER-negative regions. For regions with three MNR sets, CCV recall ranged at 72-79%, while credible set recall was reported at 91% for overall disease, 80% for ER-positive and 85% for ER-negative analysis. In all regions, credible set recall was 88.1% (88/101) for overall breast cancer, 86.4% (57/66) for ER-positive and 72% (21/29) for ER-negative, while CCV recall was 69.4% (3,823/5,510), 72.6% (898/1,238) and 52.8% (341/646) respectively.

**Fig. 3.**
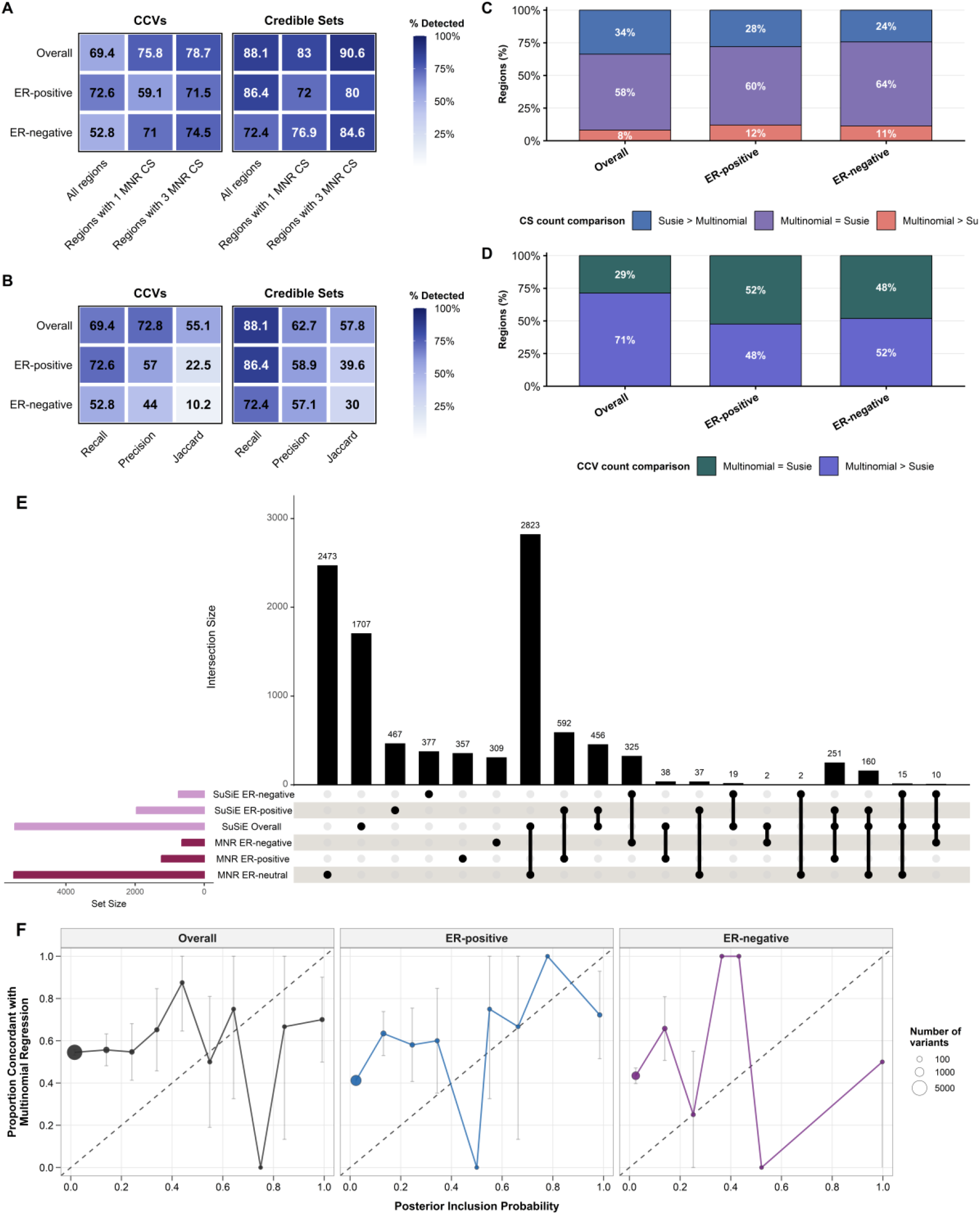
Comparison metrics, characteristics, cross-subtype overlap and PIP alignment of *SuSiE* relative to MNR. (**A**) Recall of MNR-derived CCVs and credible sets recovered by *SuSiE*, separated by phenotype and stratified according to region complexity: all 129 regions analysed, regions with a single MNR credible set (CS) and regions with three MNR CSs. (**B**) Recall, precision and Jaccard index between the two methods for CCVs and credible sets, separated by phenotype. (**C**) Among regions with at least one shared credible set, the distribution of relative credible set count is shown as follows: *SuSiE* > Multinomial (blue), equal (purple), Multinomial > *SuSiE* (orange). (**D**) Within regions with common credible sets, the proportion of credible sets with equal CCV counts (green) versus more CCVs under the multinomial model is shown. (**E**) UpSet plot summarizing CCV overlaps between phenotypes for *SuSiE* and MNR; vertical bars quantify intersection sizes, with connected dots/lines indicating shared sets and single dots indicating unique calls. (**F**) Comparison of *SuSiE* posterior inclusion probabilities with MNR CCVs across ER subtypes. Variants were binned by PIP and the proportion overlapping with multinomial CCVs was calculated per phenotype. Point size reflects the number of variants in each bin. CCV, candidate causal variant; CS, credible set; ER, estrogen receptor; MNR; Multinomial regression.

Across regions, precision and the Jaccard index were compared (**Figure 3B**). CCV precision ranged between 44-72.4% across subtypes and credible set precision ranged from 57.1-62.7%. The Jaccard index, a symmetric agreement metric where neither *SuSiE* nor MNR are considered as reference, ranged between 10.2-55.1% across phenotypes for CCVs and 30-57.8% for credible sets.

Within regions with overlapping credible sets, we compared credible set count and number of CCVs identified. The majority of regions (58-64% across phenotype analyses) had the same number of credible sets under *SuSiE* and the multinomial model (**Fig. 3C**): 24-34% had more credible sets under *SuSiE* and 8-12% had more under the multinomial model. The CCV counts in common credible sets were often equal between methods (29-52%, **Fig. 3D**); where they differed, the multinomial model more frequently included more CCVs within the shared credible sets (48-71%). No cases were observed in which *SuSiE* had produced more CCVs. Two credible sets in the multinomial analysis were deconvolved by *SuSiE* into two and three distinct credible sets respectively. The median size of all detected credible sets were larger in the multinomial analysis for overall breast cancer, but larger in the *SuSiE* analysis for ER-positive (by 1 variant) and ER-negative (by 5 variants).

MNR estimates subtype effects jointly, whereas *SuSiE* is fit separately for each phenotype using the corresponding summary statistics (overall breast cancer, ER-positive and ER-negative). To assess cross-phenotype overlap, we compared CCVs identified by phenotype-specific *SuSiE* analyses with those of the multinomial model (**Fig. 3E**). As before, MNR-defined ER-neutral CCVs were compared to *SuSiE* results using the overall breast cancer summary statistics. CCV sharing was first observed within *SuSiE* between overall breast cancer and ER-positive (n=867), whereas overlap involving the ER-negative subtype was limited (n=44). Comparison of CCVs between methods also revealed systematic differences, with some variants detected by different phenotype analyses. 38 CCVs identified by ER-positive MNR analysis were found in the overall disease analysis by *SuSiE*, while the opposite was true for 37 variants. Two variants associated to ER-neutral disease in MNR, were detected in the ER-negative analysis by *SuSiE*. In addition, *SuSiE* frequently identified MNR CCVs in multiple phenotypes, although the multinomial model assigned these to a single phenotype. For example, 160 ER-neutral CCVs and 251 ER-positive CCVs by MNR were found both to ER-positive and overall disease analyses by the summary statistics method and a total of 15 and 10 variants assigned by MNR as ER-neutral and ER-negative respectively, were found by *SuSiE* both in the overall breast cancer and ER-negative analyses.

To quantify concordance between *SuSiE* posterior inclusion probabilities (PIPs) and MNR-derived CCVs, we examined the proportion of variants within each PIP bin that overlapped multinomial CCVs (**Fig. 3F**). Overall, overlap tended to increase with higher PIP, indicating greater agreement between methods for variants assigned higher posterior probability by *SuSiE*. Aggregating across loci, MNR CCVs accounted for 58.8% of total *SuSiE* PIP in overall breast cancer, 58.9% in ER-positive, and 61.5% in ER-negative analyses, indicating that the majority of posterior mass was concentrated on variants previously prioritized by the multinomial model, while the remaining probability was distributed among variants not identified as MNR CCVs.

To further evaluate the relationship between *SuSiE* PIPs and previously reported MNR CCVs, we fitted logistic regression models with CCV overlap as the outcome and PIP as a continuous predictor. Across phenotypes, higher PIPs were significantly associated with increased probability of overlap with MNR CCVs. The association was modest but significant for overall breast cancer (β = 0.76, SE = 0.36, p = 0.031) and stronger for ER-positive (β = 3.15, SE = 0.64, p = 7.3 × 10^−7^) and ER-negative disease (β = 9.05, SE = 1.95, p = 3.4 × 10^−6^). These findings indicate that variants assigned higher posterior inclusion probabilities by *SuSiE* were increasingly enriched for CCVs identified by the multinomial model, particularly in the subtype-specific analyses.

### Investigation of Missing Credible Sets Between *SuSiE* and Multinomial Model

In total, 30 MNR-derived credible sets within 27 genomic regions, were not recovered by *SuSiE* (overall=13, ER-positive=9, ER-negative=8). Effects of *SuSiE*-nominated CCVs within these regions were explored. First, in 17 out of the 27 regions *SuSiE* did not yield any risk credible set sets or CCVs. For the remaining 10 regions, *SuSiE* identified novel credible sets not detected by the multinomial model. Although in 42% of these, *SuSiE* nominated CCVs with larger absolute effect sizes (|β|) than MNR, for the majority of them (58%), *SuSiE* tended to nominate CCVs with smaller |β| and weaker significance than those prioritized by the multinomial model. (**Supplementary Fig. 1**). Discordant credible sets were evident irrespective of the strength of the signal in these regions by MNR. Within the 10 regions the multinomial model prioritised variants with higher median |z-scores| than *SuSiE* in overall breast cancer and ER-positive variants (4.75 vs 3.38 and 5.60 vs 4.33 respectively), while for ER-negative variants, *SuSiE* prioritised variants with higher median |z-scores| than MNR (6.87 vs. 2.39). Notably, within these regions, two MNR-derived credible sets were recovered by *SuSiE* only in association to a different phenotype (a 20-variant ER-positive set and a 2-variant ER-negative set).

To assess whether CCVs in credible sets defined by *SuSiE* but not detected by the MNR might nevertheless represent the same underlying causal variants of the multinomial analysis, we evaluated linkage disequilibrium (LD) data among variants using reference data based on the European superpopulation of the 1000 Genomes Project (Phase 3). Of the 30 discordant credible sets, only five contained *SuSiE*-nominated CCVs in strong LD (r^2^ > 0.6, n = 38) with MNR-identified CCVs. This observation may reflect tagging of the same underlying causal variant from the multinomial model that was missed due to incomplete observation of the causal variant in the dataset, or method-specific differences in causal variant prioritisation. The remaining *SuSiE*-specific credible sets showed no strong correlation with MNR CCVs, potentially representing novel sets or refined localizations unique to *SuSiE*.

We next explored whether genotype discordance due to allele flipping could explain the missing credible sets using *SuSiE’s* diagnostic plots of observed versus expected z-scores. We compared diagnostic plots for 11 regions in which not all multinomial credible sets were detected by *SuSiE* (**Supplementary Fig. 2**) and examples of regions in which all credible sets were detected by both methods (**Supplementary Fig. 3**). Although evidence of potential allele flips was observed in all regions; their frequency did not differ between regions with overlapping sets and those with missing credible sets, indicating that allele flipping is unlikely to explain the discordance.

To assess whether data quality issues in the summary statistics could explain the missing credible sets, we annotated the 129 genomic regions with SLALOM, a quality control tool that detects heterogeneous or outlier summary statistics in meta-analysis fine-mapping based on consistency with markers in LD. Of the 27 regions in which *SuSiE* and MNR credible sets were discordant, four (∼15%) were flagged as suspicious by SLALOM, including one region for overall/ER-negative disease (chr5:57684061-58865569) and three regions for ER-positive disease (chr1:154648781-155648781; chr11:68831418-69879161; chr2:171876221-173472971), indicating harmonization-related heterogeneity and potentially unstable fine-mapping.

Further to these annotations, we examined whether imputation quality measured by INFO scores for variants in the 27 discordant regions could be influencing fine-mapping accuracy (**Supplementary Fig. 4**). Imputation quality was generally lower for iCOGS than for OncoArray, with pronounced differences in a subset of regions (e.g., chr1:145144984-146144984). Nevertheless, 75% of discordant regions had mean INFO ≥ 0.80, indicating that array-level imputation quality is unlikely to be the primary driver of the observed discordance. When comparing the distribution of average imputation scores between iCOGS and OncoArray, between MNR-derived CCVs within missing credible sets and MNR-derived CCVs within overlapping credible sets (**Supplementary Fig. 5**), imputation score distributions differed (Wilcoxon rank-sum test, BH-adjusted p < 10^−3^), with overlapping CCVs showing consistently higher scores. Effect sizes, quantified as rank-biserial correlations from the Wilcoxon rank-sum test, were small for overall breast cancer and ER-positive disease (0.11-0.20) but moderate for ER-negative disease (0.49), indicating that overlapping CCVs tend to have higher imputation scores than missing CCVs, particularly in ER-negative loci. When comparing the iCOGS or OncoArray imputation scores for the two CCV groups, only minimal differences were observed.

To further assess potential differences in *SuSiE* results across genotyping arrays, we repeated the fine-mapping analyses using summary statistics from iCOGS and OncoArray separately. The meta-analysis summary statistics showed the highest MNR-derived credible set recall by *SuSiE*. However, in the overall breast cancer OncoArray summary statistics analysis, four additional credible sets, previously missing, were recovered in two genomic regions (one of these was also recovered in the iCOGS-based analysis). As a result, credible set recall increased from 88.1% to 90% in the overall disease analysis. Absolute z-score distributions of variants within the recovered credible sets were weaker in the OncoArray compared to the meta-analysis.

To further assess unambiguous credible sets, we focused on MNR credible sets containing a single CCV (n=28). These single-CCV sets (‘singleton sets’) represent association sets without additional LD-correlated variants, removing ambiguity caused by multi-variant sets and providing a clear benchmark. CCV and credible set recall was reported at 85.7% (n=24/28). For the missing CCVs, imputation quality was between 0.83-0.99 except for one variant with a score of 0.36 in iCOGS and 0.45 in OncoArray. No missing CCVs from singleton variants were in strong LD with *SuSiE*-specific credible sets and CCVs.

### Impact of LD source on *SuSiE* fine-mapping

We evaluated the impact of the LD matrix source on fine-mapping by comparing results using in-sample LD matrices from OncoArray versus out-of-sample matrices derived from the UK Biobank. As before, results were compared to the MNR model using, for each region, the value of L that maximised recall with the multinomial stepwise regression (**Fig. 4A**). Using the UK Biobank matrices, *SuSiE* detected 95 additional credible sets were detected, compared to the in-sample analysis. While the median number of credible sets per region remained consistent between analyses for overall breast cancer (1.0) and ER-negative (1.0), it increased from 1.0 to 2.0 for ER-positive using the UK Biobank matrices. Notably, the median credible set size decreased substantially with out-of-sample LD matrices: by 92% for overall breast cancer (from 13 to 1 variant; Wilcoxon rank-sum P = 4.0 × 10^−11^), 73% for ER-positive disease (from 11 to 3; P = 3.4 × 10^−4^) and 26% for ER-negative disease (from 13.5 to 10; p = 0.36).

**Fig. 4.**
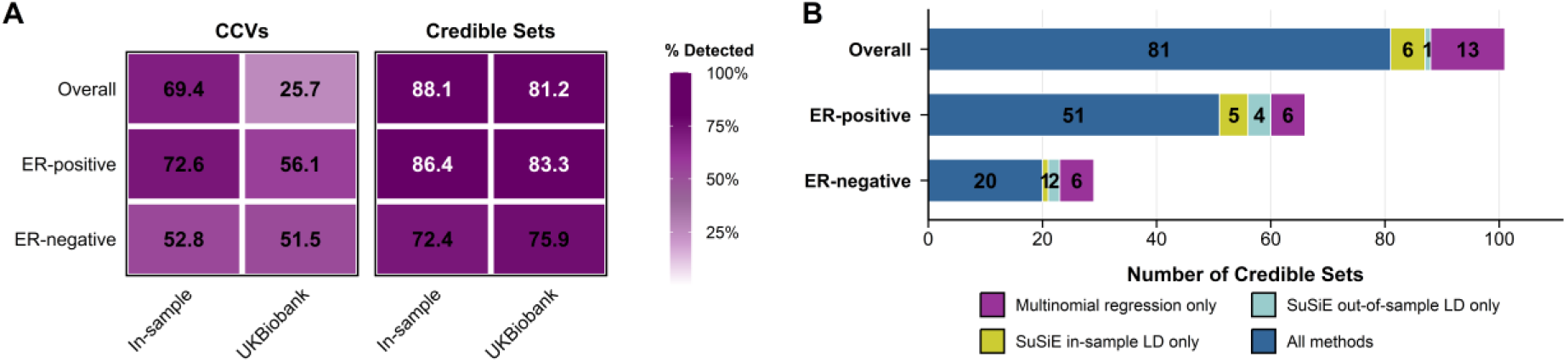
*SuSiE* recall using in-sample versus out-of-sample LD matrices and MNR-derived credible sets. (**A**) Recall of MNR-derived CCVs and credible sets recovered by *SuSiE* using in-sample LD matrices vs out-of-sample LD matrices (UK Biobank), separated by phenotype. Recall is calculated for all 129 regions analysed. (**B**) Stacked bar chart showing the number of MNR-derived credible sets (Fachal *et al*. 2020) detected by *SuSiE* using in-sample versus out-of-sample (UK Biobank) LD matrices across phenotypes. Numbers within bars indicate the count of credible sets in each category.

CCV recall was substantially reduced when using out-of-sample matrices for overall disease (from 54% to 26%) and ER-positive (from 68% to 56%) subtypes, while ER-negative recall remained comparable between analyses. Compared to the in-sample LD analysis, credible set recall using out-of-sample LD matrices showed modest decline for overall breast cancer (from 88.1% to 81%) and ER-positive (from 86.4% to 83%), while ER-negative showed a slight improvement (from 72.4% to 76%). For most MNR-credible sets, only a few CCVs were recovered using out-of-sample matrices: the median number of CCVs recalled per MNR-credible set was 1, 6, and 7 for overall disease, ER-positive, and ER-negative subtypes, respectively, explaining the low CCV recall rates observed, especially in the case of overall disease. When considering all phenotypes, CCV and credible set recall dropped by 23% and 3% using out-of-sample matrices compared to the in-sample analysis.

A total of 37 MNR-derived credible sets were discordant using external LD reference, compared to 30 using in-sample LDs. Importantly, the discordant sets differed between the two LD analyses, where the additional undetected sets with the external LD were not identical to those missed with in-sample LD **Fig. 4B)**. Specifically, 25 unique credible sets were identified exclusively by MNR, 12 by *SuSiE* in-sample and 7 by *SuSiE* out-of-sample analysis.

### Gene target annotation of *SuSiE* predictions

*SuSiE* using in-sample LD matrices identified 5,481 CCVs for overall breast cancer, 1,963 for the ER-positive subtype and 748 for the ER-negative subtype. Out of these, 42% of CCVs in all phenotypes were considered new (i.e., not detected by MNR) and corresponded to 117 novel credible sets. The INQUISIT pipeline was used to determine the likely gene targets of all *SuSiE*-derived CCVs, whether in MNR or not, categorised as coding, distal and promoter predictions. *SuSiE*-predicted CCVs were mapped to 252 level 1 (high confidence) gene targets, including 18 cancer drivers. For the overall breast cancer credible sets, the recall rate of genes annotated to *SuSiE*-derived CCVs relative to MNR-derived predictions was 92%. By subtype, 74% of ER-positive and 63% of ER-negative predictions were recalled by *SuSiE*. Thus, a total of 52 genes annotated to multinomial-derived CCVs were absent from the *SuSiE*-based INQUISIT analysis (**Supplementary Table S3 & S4**), but 42% of these were identified at a lower confidence level (level 2 in the pipeline). 32 out of the 52 missing targets corresponded to discordant credible sets between *SuSiE* and MNR and included the known breast cancer driver genes *WNT7B, CDKAL1, CREBBP* and *CCNE1*. Any remaining genes missing from the *SuSiE*-based analysis mapped to credible sets found by both methods, which likely reflects either CCVs that were not consistently detected between the two methods despite at least one overlapping CCV (for each overlapping credible set), or differences in the data sources used in the original INQUISIT pipeline. Finally, 30 new genes (level 1) were identified in the *SuSiE*-based analysis including three known breast cancer drivers *KANSL1, CACNA1A, ZNF703* and other breast cancer associated genes such as *NEK10, ZNF575, GATA3-AS1, MAPT, EZH1*.

### Assessment of fine-mapping results with functionally-characterized variants

A systematic literature review from 25 published breast cancer fine-mapping studies was performed to explore available experimental functional evidence for (i) new CCVs detected for the first time in *SuSiE* credible sets not found by MNR and (ii) MNR CCVs that were missed by *SuSiE*. In 16 of the 25 published studies, at least one likely causal variant had been experimentally validated. In region chr4:105569013-106856761 (TET2), *SuSiE* recovered the full multinomial credible set (100%), although the index CCV differed: *SuSiE* prioritized rs62331150, previously validated to affect active promoter and enhancer regions of TET2 in human mammary epithelial cells, whereas the multinomial index variant lacked functional support. At chr5:55531884-56587883 (MAP3K1), *SuSiE* captured all multinomial credible sets and identified rs16886397 and rs17432750, both confirmed by luciferase assays to impact MAP3K1 promoter activity and GATA3 binding; however, *SuSiE* missed two multinomial CCVs (rs832402, rs7721581) identified as SETD9 eQTLs in TCGA and METABRIC datasets. In the 6q25 region (chr6:151418856-152937016; ESR1), *SuSiE* detected 5 of 6 multinomial credible sets and identified an additional credible set containing rs2747652 and rs910416, variants shown to influence MYC transcription factor binding and increase risk for ER^−^PR^−^HER2+ tumours. Conversely, at chr11:68831418-69879161, *SuSiE* recovered 67% of multinomial credible sets; the multinomial index included rs661204, experimentally shown to reduce CUPID1 enhancer activity and abrogate chromatin looping, whereas the *SuSiE* lead SNP, rs72927602, lacked functional evidence. RegulomeDB annotation of *SuSiE*-specific CCVs at this locus revealed generally modest regulatory potential, with most variants affecting transcription factor binding in non-breast tissues or inactive chromatin states, except rs655786, which localizes to active breast enhancers and shows enrichment for multiple transcription factor binding sites in mammary epithelium.

## Discussion

Fine-mapping attempts to determine the variants at GWAS signals with a direct effect on a trait. Multiple statistical approaches have been developed with different features and capabilities, such as the incorporation of summary statistics. However, most of the tools have been evaluated on simulated datasets and quantitative traits. To date, no detailed comparison of methods and findings has been performed on cancer traits, leaving a significant gap in assessing the real-world applicability and performance of the fine-mapping methods.

In breast cancer which is one of the largest GWAS resources of any complex trait and a large number of loci has been identified, a previous multinomial fine-mapping analysis using the conventional stepwise regression approach identified 196 strong independent causal sets and 191 likely target genes^[41]^. This study represents the most comprehensive collection of causal variants in breast cancer. Here, we aimed to compare results generated by *SuSiE*, one of the most commonly-used Bayesian summary statistics methodologies, against these published results derived by multinomial regression using individual-level data on the same meta-analysis dataset. *SuSiE* enables the detection of multiple independent causal variants per region at high computational efficiency, either from individual or summary statistics data^[7,20]^. Two other summary statistics methods, *finiMOM* and *FINEMAP* were also assessed, however *SuSiE* was selected for comprehensive comparison to the multinomial model, having the highest recall and lowest CCVs inflation.

A key user-defined parameter in *SuSiE* is the maximum number of independent causal variants allowed per region (L), whereas in the multinomial stepwise model this number is implicitly determined by the stepwise selection procedure. Our evaluation of the optimal L parameter revealed that CCV and credible set recall as well as the number of regions producing causal sets generally declined as the assumed number of causal variants increased, although previous work suggests that overestimating L is generally robust^[7,20]^ and a common practice aiming to capture all potential causal sets. Furthermore, there was considerable heterogeneity across regions: some regions lost credible sets at higher L values, while others only began to yield results under these assumptions. These findings indicate that regions do not respond uniformly to changes in L and that applying a single universal L parameter across regions may lead to the omission of important credible sets. We therefore recommend evaluating the choice of the maximum number of causal variants per region, particularly at higher values of L. One potential strategy to mitigate this issue when applying *SuSiE* is to evaluate each region across a range of L values, particularly for regions where no credible sets are initially detected, and select the specification that yields stable and informative results, such as consistent CCV and/or credible set recovery or the identification of high-purity credible sets.

As the primary objective of the study was to compare *SuSiE* results with previously published multinomial regression fine-mapping results, we selected, for each region, the L value that maximized recall relative to the multinomial stepwise approach. Based on these settings, *SuSiE* identified 139 credible sets (5,481 CCVs) in the overall breast cancer analysis, 81 in the ER-positive subtype analysis (1,963 CCVs) and 24 sets (748 CCVs) in the ER-negative analysis. Compared to MNR, *SuSiE* identified more credible sets in 24-34% of regions and nominated more CCVs in 35-47% of regions. Conversely, the multinomial approach identified more credible sets than *SuSiE* in 8-12% of regions and more CCVs in 30-41% of regions, indicating that neither method consistently outperforms the other across regions. When comparing credible set recall to MNR, reasonable agreement was observed, with 88% credible set recall in overall breast cancer, 86% in ER-positive and 72% for ER-negative subtype. However, CCV recall by *SuSiE* was much poorer at 69% for overall disease, 73% for ER-positive and 53% for ER-negative subtype, in line with the substantial detection of new CCVs by the method (at 87% of total CCVs detected). Interestingly within concordant credible sets, found by both methods, CCV count was either higher in MNR (52-71%) or of equal number (29-52%). The Jaccard index, assessing relatedness in the fine-mapping approaches without treating either as a reference set, ranged between 10.2-55.1% across phenotypes for CCVs and 30-57.8% for credible sets, which can be explained by the substantial method-specific variant nomination. For example, in overall breast cancer, although 3,824 CCVs were shared between the methods, we observe 1,430 variants identified only by *SuSiE* and 1,686 variants only by MNR. Similar evidence was observed for ER-positive and ER-negative.

Using the same strategy of selecting the fine-mapping results that maximized recall relative to the multinomial stepwise approach, *SuSiE* was additionally applied using external reference LD data derived from British ancestry individuals in the UK Biobank. This comparison was performed to reflect common practice in summary-statistics-based fine-mapping, where in-sample LD matrices are often unavailable^[42]^. Overall, the choice of LD reference panel influenced fine-mapping results. Relative to in-sample LD, use of out-of-sample matrices increased the number of detected credible sets and reduced credible set size across subtypes. However, this was accompanied by a marked reduction in CCV recall (to 26.7% for overall disease, 56.1% in ER-positive disease and 51.5% in ER-negative disease). At the credible set level, only minimal differences were observed using the external reference. Comparison with MNR further revealed shifts in concordance patterns: some sets overlapped with the multinomial model only under in-sample LD, whereas others were exclusively detected under the out-of-sample LD. These apparent gains in credible sets likely represent false positive results arising from ancestry mismatch between summary statistics and LD^[21]^, emphasizing the importance of using in-sample LD for stability and reliability of fine-mapping.

Overall, in the in-sample LD analysis, 30 credible sets in 27 genomic regions from the MNR analysis did not overlap any credible sets from the *SuSiE* analyses. Discordant credible sets were observed even for regions with strong association signals in MNR. Of these, 17 regions produced no credible sets in *SuSiE*, while the remaining 10 produced novel sets not detected by the multinomial. Several factors potentially influencing differences in fine-mapping results were explored. First, LD analysis showed that 38 variants in five *SuSiE*-specific credible sets are in strong correlation (r^2^ > 0.6) with multinomial-identified CCVs. It is therefore possible that the novel sets represent the same causal variant sets, with one or other method incorrectly excluding the causal variants due to methodological differences (or potentially because the causal variants is not represented in the dataset). The remaining *SuSiE*-specific sets showing no LD correlation with multinomial CCVs may represent secondary signals with weaker effects better captured by *SuSiE’s* iterative framework, or spurious associations from model misspecification. Distinguishing between these requires functional validation.

We also explored the possibility of allele flips between summary statistics and in-sample LD matrices as a potential source of discordant credible sets. Of the 27 regions with discordant sets, five showed allele flips in at least one CCV. However, allele flipping of multinomial-nominated CCVs did not appear to be related to discordance. Moreover, four of the 27 discordant regions were flagged as suspicious by SLALOM, representing regions in which the meta-analysis summary statistics data used in this study and potential heterogeneity within them, may be leading to reduced fine-mapping accuracy. These were significantly depleted for likely causal variants among high-PIP variants^[21]^. In continuation of these findings, imputation quality between arrays was analysed for the discordant regions. Imputation quality was generally lower for iCOGS than the OncoArray, however all discordant regions exceeded INFO > 0.30 and 75% had mean INFO > 0.80. More importantly, distributional differences of imputation scores between MNR-derived CCVs within discordant credible sets and MNR-derived CCVs within overlapping credible sets differed (Wilcoxon rank-sum test, BH-adjusted p < 10^−3^), with overlapping CCVs showing consistently higher scores. To account whether imputation quality between arrays or other types of array heterogeneity may be contributing to fine-mapping accuracy, *SuSiE* was applied separately on summary statistics from iCOGS and OncoArray which improved overall breast cancer credible set concordance to 92% by recovering four credible sets using the OncoArray summary statistics alone. Absolute z-score distributions of variants within the recovered credible sets were weaker in the OncoArray compared to the meta-analysis. One credible set was recovered by both iCOGS- and OncoArray-based analyses but not using the meta-analysis data.

Although the above findings indicate that array heterogeneity, imputation quality and allele flips may be influencing fine-mapping, these alone do not explain the degree of discordance observed. Within the regions that *SuSiE* produced results but no overlaps with MNR were detected (n=17), we find that the majority of sets (68%) nominate CCVs with smaller |β| and weaker significance than those prioritized by the multinomial model. Therefore, method-specific differences between the two pipelines, including effect-size weighting could be the reason behind discordant results. This raises questions about whether these events represent genuine refinements or reflect *SuSiE’s* more liberal credible set construction when L exceeds the true number of causal variants. Further to that, when assessing recall within singleton sets, i.e., MNR-derived credible sets with a single CCV, *SuSiE* reached 86% (n=24/28), suggesting that the *SuSiE* reliably recovers most of the MNR-identified credible sets. Overall, recall was higher for unambiguous, independent association credible sets, whereas lower concordance in regions harbouring multiple independent causal variants likely reflects the inherently difficult challenges of resolving correlated variants. Moreover, the two methods differ fundamentally in their modelling frameworks; the classical implementation of *SuSiE* evaluates associations with each phenotype separately, while MNR models phenotypes jointly. Given that breast cancer is highly heterogeneous, it is possible that multiple univariate regression does not perform as well as multivariate regression. In addition, as a consequence of this fundamental difference in phenotype modelling, the filtering criteria applied in the original MNR analysis by Fachal et al., (2020) such as MAF and P value, were required to be met for at least one of the three phenotypes (overall disease, ER-positive and ER-negative). This approach was not directly transferrable to *SuSiE*, in which identical filtering thresholds were applied independently to each phenotype. A secondary consequence of this methodological distinction is that the effective sample size and statistical power differ across phenotypes, which may further contribute to differences in CCV overlap between methods. For example, it was observed that the CCVs in ER-negative credible sets in *SuSiE* had substantially stronger associations that those in MNR credible sets in the same region, consistent with the fact that *SuSiE* analyses had less power to detect ER-negative associations. Extension of Bayesian summary statistics methods to multivariate endpoints might rectify this.

Although a primary goal of this study was to compare the MNR framework to summary-statistics-based fine-mapping by *SuSiE* – a widely applied method – we acknowledge that the use of summary statistics in *SuSiE*, as opposed to individual-level genotype data in the MNR analysis may further contribute to differences in fine-mapping accuracy. We also acknowledge the following caveats that may have contributed to discordant results. First, in the *SuSiE* analysis multi-allelic variants were excluded to minimise potential distortion in fine-mapping, a filtering step not applied in the original MNR analysis by Fachal et al., (2020). A sensitivity analysis was conducted in regions harbouring multi-allelic variants using the same input variant set as the original MNR analysis, and no differences were observed in credible set or CCV recall. In addition, although the summary statistics used in the *SuSiE* analysis were derived from a meta-analysis of iCOGS and OncoArray, the in-sample LD data used were computed using the OncoArray data only, given the practical challenge of generating LD matrices jointly across array platforms. As fine-mapping is sensitive to mismatches between genotyping arrays and imputation panels^[21]^, this discrepancy may have influenced calibration and downstream results. By contrast, the MNR framework derived LD matrices separately for each dataset, representing an additional fundamental methodological difference between *SuSiE* and MNR. These factors were addressed in the array-specific sensitivity analyses, which may also explain the recovery of four previously discordant sets in the OncoArray-restricted analysis.

In this study, recall was used as the primary comparison metric to MNR. Although the multinomial regression approach reported by Fachal et al. cannot be considered a ground truth, it currently represents the most comprehensive fine-mapping evaluation and curated set of candidate causal variants for breast cancer. Thus, comparing alternative approaches against this framework informs the future application of *SuSiE* in fine-mapping.

The absence of ground truth in real data with which to compare to represents a fundamental challenge in the evaluation of fine-mapping methods, and underlies the use of simulated datasets to assess their performance. One commonly used approach is to leverage orthogonal functional data, but this is difficult to apply systematically because only a limited number of regions have been experimentally interrogated, various techniques have been used and variants are often preselected based on prior association evidence. In our study, we identified 16 regions in which likely causal variants has been experimentally characterised. For example, at the *TET2* locus (chr4:105569013-106856761), *SuSiE* recovered the full multinomial credible set, although the top-ranked variant differed: rs62331150 has been validated to affect promoter and enhancer activity in human mammary epithelial cells, whereas the multinomial index variant lacked functional evidence^[43]^. The inclusion of experimentally supported variants within the credible set illustrates that both methods capture biologically relevant signals. Similarly, at the *MAP3K1* locus (chr5:55531884-56587883), *SuSiE* captured all multinomial credible sets and included rs16886397 and rs17432750, both functionally confirmed to influence promoter activity and GATA3 binding ^[36]^. However, two multinomial CCVs were missed by *SuSiE* were previously found to act as *SETD9* eQTLs in TCGA and METABRIC datasets. At the *ESR1* locus (chr6:151418856-152937016), *SuSiE* recovered 5 out of 6 multinomial credible sets and identified a new credible set not captured by the multinomial model, that were previously found to modulate MYC binding and to significantly increase risk for ER^−^PR^−^HER2+ tumours responsive to trastuzumab^[37]^. Luciferase assays also confirmed these variants enhance promoter activity in both ER+ and ER-breast cell lines. Conversely, at chr11:68831418-69879161, *SuSiE* recovered only two of three MNR credible sets, missing the experimentally validated rs661204 found to reduce *CUPID1* enhancer activity ^[49]^, while the *SuSiE*-specific lead SNP rs72927602 lacks functional evidence. RegulomeDB annotation of *SuSiE*-specific CCVs at this locus indicated generally modest regulatory potential, with most variants affecting non-breast tissues or inactive chromatin, except rs655786, which maps to active enhancers and shows multiple transcription factor binding events in mammary epithelium^[44]^. Collectively, these examples illustrate that while complementary fine-mapping methods can capture biologically relevant signals, there is substantial heterogeneity in which variants are prioritized. Although functional evidence alone is not definitive, integrating complementary fine-mapping approaches with orthogonal functional data can aid the interpretation of association signals, taking both methodological and experimental limitations into account.

The biological significance of identified CCVs and credible sets was also explored by target gene annotation using INQUISIT. When compared to MNR-based INQUISIT analysis, 52 genes including established breast cancer drivers such as *CCNE1* and *BRCA2* were absent from the *SuSiE*-based INQUISIT analysis; although some of these were captured at lower confidence (level 2 in the pipeline). Out of the 52 missing genes, 32 corresponded to discordant credible sets between the two methods. The remaining missing genes corresponded to overlapping credible sets, reflecting either inconsistent CCVs detected by the two methods despite at least one overlapping CCV (for each overlapping credible set), or differences in the data sources used in the original INQUISIT pipeline, such as variation in tools for annotating splicing or missense variants. Importantly, *SuSiE* identified 3,440 new variants (not detected by MNR), leading to the annotation of 30 new level 1 genes. Three of these are the known breast cancer drivers *KANSL1, CACNA1A, ZNF703* as well as breast cancer associated genes such as *NEK10, ZNF575, GATA3-AS1, MAPT, EZH1*. These novel findings suggest that integrating predictions from complementary fine-mapping approaches may provide a more reliable set of gene targets than relying on a single method.

In conclusion, this comparative analysis provides insights for future fine-mapping studies and demonstrates that the results of fine-mapping analyses can differ substantially between methods and should not be interpreted without critical evaluation. Multivariate regression methods and Bayesian summary statistics approaches such as *SuSiE* have strengths and weaknesses, indicating that interpretations should be made carefully. Regional complexity, including intricate linkage disequilibrium architecture, polygenicity and the presence of multiple independent causal variants and credible sets within loci, impacts both method performance and inter-method concordance rates. Additional factors contributing to discordant results include array-based heterogeneity and genetic heterogeneity across breast cancer subtypes. These observations emphasize that validation through orthogonal fine-mapping algorithms, combined with functional assays including luciferase reporter assays and eQTL analyses, as well as integration with regulatory genomic annotations such as chromatin state and transcription factor binding profiles, is essential for confident causal variant prioritization.

## Data Availability

All data produced in the present work are contained in the manuscript

## Code availability

The *SuSiE* software is available at https://github.com/stephenslab/susieR. *LDstore2* and *FINEMAP* are available at http://www.christianbenner.com/. *finiMOM* is available at https://vkarhune.github.io/finimom/.

## Acknowledgements

Joe Dennis is supported by core funding from the NIHR Cambridge Biomedical Research Centre (NIHR203312).

## References

1. Michailidou K, Lindström S, Dennis J, Beesley J, Hui S, Kar S, et al. Association analysis identifies 65 new breast cancer risk loci. 2017; Nature.

2. Zhang H, Ahearn TU, Lecarpentier J, Barnes D, Beesley J, Qi G, et al. Genome-wide association study identifies 32 novel breast cancer susceptibility loci from overall and subtype-specific analyses. 2020; Nat Genet.

3. Fachal L, Aschard H, Beesley J, Barnes DR, Allen J, Kar S, et al. Fine-mapping of 150 breast cancer risk regions identifies 191 likely target genes. 2020; Nat Genet.

4. Spain SL, Barrett JC. Strategies for fine-mapping complex traits. 2015; Hum Mol Genet.

5. Edwards SL, Beesley J, French JD, Dunning AM. Beyond GWASs: Illuminating the Dark Road from Association to Function. 2013; Am J Hum Genet.

6. Schaid DJ, Chen W, Larson NB. From genome-wide associations to candidate causal variants by statistical fine-mapping. 2018; Nat Rev Genet.

7. Zou Y, Carbonetto P, Wang G, Stephens M. Fine-mapping from summary data with the “Sum of Single Effects” model. 2022; PLoS Genet.

8. Zhang W, Najafabadi H, Li Y. SparsePro: An efficient fine-mapping method integrating summary statistics and functional annotations. 2023; PLOS Genet.

9. Xu S, Williams J, Tegge A, Ferreira MAR. Genome-wide iterative fine-mapping for non-Gaussian phenotypes. 2025; Sci Reports 2025 151.

10. Akdeniz BC, Frei O, Shadrin A, Vetrov D, Kropotov D, Hovig E, et al. Finemap-MiXeR: A variational Bayesian approach for genetic finemapping. 2024; PLOS Genet.

11. Karhunen V, Launonen I, Järvelin MR, Sebert S, Sillanpää MJ. Genetic fine-mapping from summary data using a nonlocal prior improves the detection of multiple causal variants. 2023; Bioinformatics.

12. Wu Y, Zheng Z, Thibaut L, Lin T, Feng Q, Cheng H, et al. Genome-wide fine-mapping improves identification of causal variants. 2025; medRxiv.

13. Weissbrod O, Hormozdiari F, Benner C, Cui R, Ulirsch J, Gazal S, et al. Functionally informed fine-mapping and polygenic localization of complex trait heritability. 2020; Nat Genet.

14. Shrestha M, Bai Z, Gholipourshahraki T, Hjelholt AJ, Hu S, Kjolby M, et al. Enhanced genetic fine mapping accuracy with Bayesian Linear Regression models in diverse genetic architectures. 2025; PLOS Genet.

15. Karhunen V, Launonen I, Järvelin MR, Sebert S, Sillanpää MJ. Genetic fine-mapping from summary data using a nonlocal prior improves the detection of multiple causal variants. 2023; Bioinformatics.

16. Amos CI, Dennis J, Wang Z, Byun J, Schumacher FR, Gayther SA, et al. The OncoArray Consortium: A Network for Understanding the Genetic Architecture of Common Cancers. 2017; Cancer Epidemiol Biomarkers Prev.

17. Michailidou K, Hall P, Gonzalez-Neira A, Ghoussaini M, Dennis J, Milne RL, et al. Large-scale genotyping identifies 41 new loci associated with breast cancer risk. 2013; Nat Genet.

18. Benner C, Havulinna AS, Järvelin MR, Salomaa V, Ripatti S, Pirinen M. Prospects of Fine-Mapping Trait-Associated Genomic Regions by Using Summary Statistics from Genome-wide Association Studies. 2017; Am J Hum Genet.

19. Benner C, Spencer CCA, Havulinna AS, Salomaa V, Ripatti S, Pirinen M. FINEMAP: Efficient variable selection using summary data from genome-wide association studies. 2016; Bioinformatics.

20. Wang G, Sarkar A, Carbonetto P, Stephens M. A simple new approach to variable selection in regression, with application to genetic fine mapping. 2020; J R Stat Soc Ser B Stat Methodol.

21. Kanai M, Elzur R, Zhou W, Wu KHH, Rasheed H, Tsuo K, et al. Meta-analysis fine-mapping is often miscalibrated at single-variant resolution. 2022; Cell Genomics.

22. Corradin O, Saiakhova A, Akhtar-Zaidi B, Myeroff L, Willis J, Cowper-Sallari R, et al. Combinatorial effects of multiple enhancer variants in linkage disequilibrium dictate levels of gene expression to confer susceptibility to common traits. 2014; Genome Res.

23. He B, Chen C, Teng L, Tan K. Global view of enhancer-promoter interactome in human cells. 2014; Proc Natl Acad Sci U S A.

24. Andersson R, Gebhard C, Miguel-Escalada I, Hoof I, Bornholdt J, Boyd M, et al. An atlas of active enhancers across human cell types and tissues. 2014; Nature.

25. Hnisz D, Abraham BJ, Lee TI, Lau A, Saint-André V, Sigova AA, et al. Transcriptional super-enhancers connected to cell identity and disease. 2013; Cell.

26. Weinstein JN, Collisson EA, Mills GB, Shaw KRM, Ozenberger BA, Ellrott K, et al. The Cancer Genome Atlas Pan-Cancer analysis project. 2013; Nat Genet.

27. Curtis C, Shah SP, Chin SF, Turashvili G, Rueda OM, Dunning MJ, et al. The genomic and transcriptomic architecture of 2,000 breast tumours reveals novel subgroups. 2012; Nat 2012 4867403.

28. Dixon JR, Xu J, Dileep V, Zhan Y, Song F, Le VT, et al. Integrative detection and analysis of structural variation in cancer genomes. 2018; Nat Genet.

29. Dunham I, Kundaje A, Aldred SF, Collins PJ, Davis CA, Doyle F, et al. An integrated encyclopedia of DNA elements in the human genome. 2012; Nature.

30. Hammal F, De Langen P, Bergon A, Lopez F, Ballester B. ReMap 2022: a database of Human, Mouse, Drosophila and Arabidopsis regulatory regions from an integrative analysis of DNA-binding sequencing experiments. 2022; Nucleic Acids Res.

31. McLaren W, Gil L, Hunt SE, Riat HS, Ritchie GRS, Thormann A, et al. The Ensembl Variant Effect Predictor. 2016; Genome Biol.

32. Jaganathan K, Kyriazopoulou Panagiotopoulou S, McRae JF, Darbandi SF, Knowles D, Li YI, et al. Predicting Splicing from Primary Sequence with Deep Learning. 2019; Cell.

33. Shamsani J, Kazakoff SH, Armean IM, McLaren W, Parsons MT, Thompson BA, et al. A plugin for the Ensembl Variant Effect Predictor that uses MaxEntScan to predict variant spliceogenicity. 2018; Bioinformatics.

34. Jian X, Boerwinkle E, Liu X. In silico prediction of splice-altering single nucleotide variants in the human genome. 2014; Nucleic Acids Res.

35. Adzhubei I, Jordan DM, Sunyaev SR. Predicting Functional Effect of Human Missense Mutations Using PolyPhen-2. 2013; Curr Protoc Hum Genet.

36. Ng PC, Henikoff S. SIFT: predicting amino acid changes that affect protein function. 2003; Nucleic Acids Res.

37. Ioannidis NM, Rothstein JH, Pejaver V, Middha S, McDonnell SK, Baheti S, et al. REVEL: An Ensemble Method for Predicting the Pathogenicity of Rare Missense Variants. 2016; Am J Hum Genet.

38. Liu X, Li C, Mou C, Dong Y, Tu Y. dbNSFP v4: a comprehensive database of transcript-specific functional predictions and annotations for human nonsynonymous and splice-site SNVs. 2020; Genome Med.

39. Peng D, Li H, Hu B, Zhang H, Chen L, Lin S, et al. PTMsnp: A Web Server for the Identification of Driver Mutations That Affect Protein Post-translational Modification. 2020; Front Cell Dev Biol.

40. Boyle AP, Hong EL, Hariharan M, Cheng Y, Schaub MA, Kasowski M, et al. Annotation of functional variation in personal genomes using RegulomeDB. 2012; Genome Res.

41. Fachal L, Aschard H, Beesley J, Barnes DR, Allen J, Kar S, et al. Fine-mapping of 150 breast cancer risk regions identifies 191 likely target genes. 2020; Nat Genet.

42. Pasaniuc B, Price AL. Dissecting the genetics of complex traits using summary association statistics. 2016; Nat Rev Genet 2016 182.

43. Guo X, Long J, Zeng C, Michailidou K, Ghoussaini M, Bolla MK, et al. Fine-scale mapping of the 4q24 locus identifies two independent loci associated with breast cancer risk. 2015; Cancer Epidemiol Biomarkers Prev.

44. Dunning AM, Michailidou K, Kuchenbaecker KB, Thompson D, French JD, Beesley J, et al. Breast cancer risk variants at 6q25 display different phenotype associations and regulate ESR1, RMND1 and CCDC170. 2016; Nat Genet.

