## Supplementary Material for "Comparative fine-mapping of breast cancer susceptibility loci using summary statistics methods and multinomial regression"

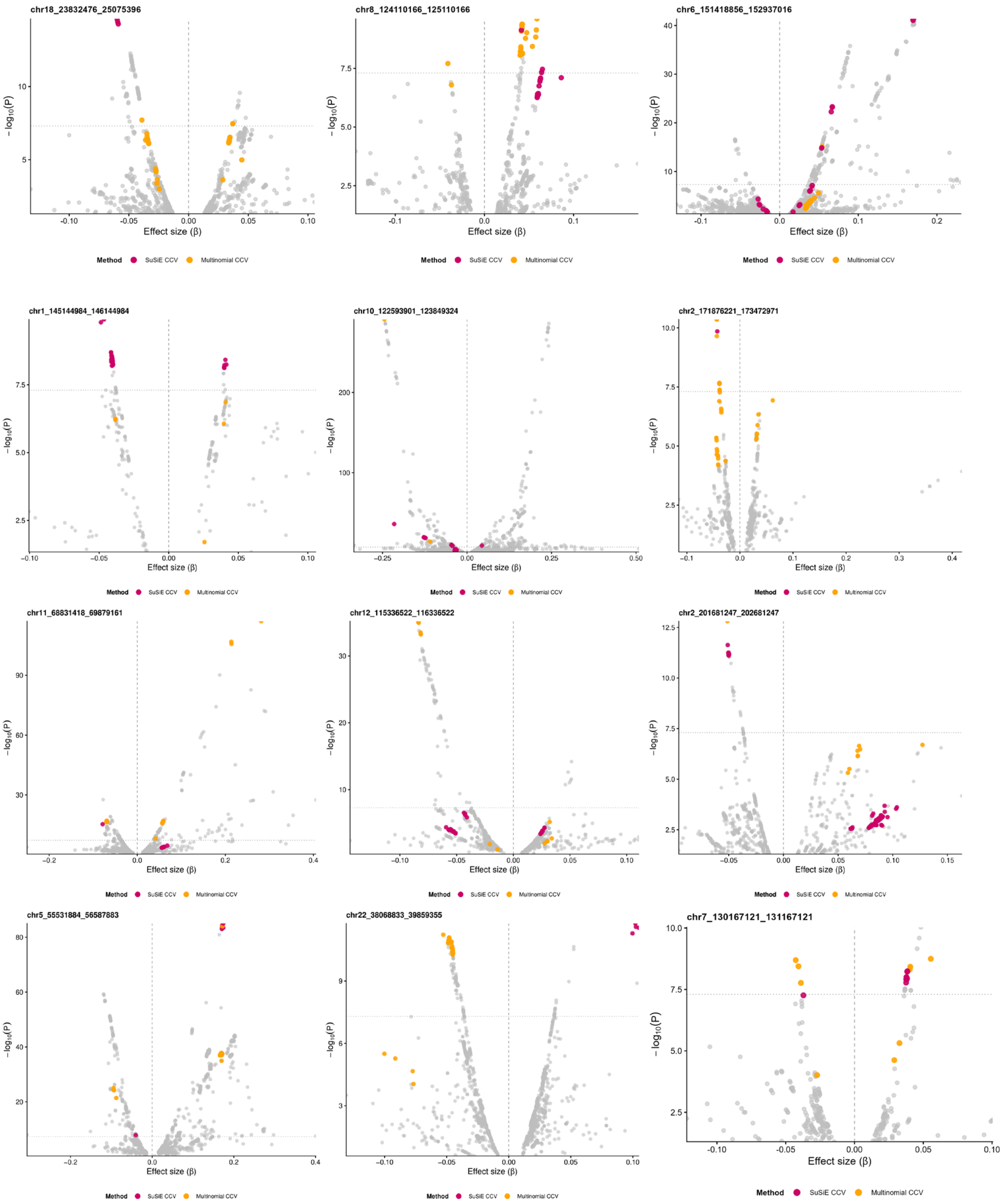


**Supplementary Figure 1. Effect size and significance of variants within regions with discordant credible sets between *SuSiE* and Multinomial.** Coloured points highlight candidate causal variants (CCVs): SuSiE CCVs (magenta) and multinomial CCVs (orange). Grey points are all other variants in the region. The vertical dashed line marks β = 0; the horizontal dotted line marks genome-wide significance ($P = 5\times10^{-8}$).

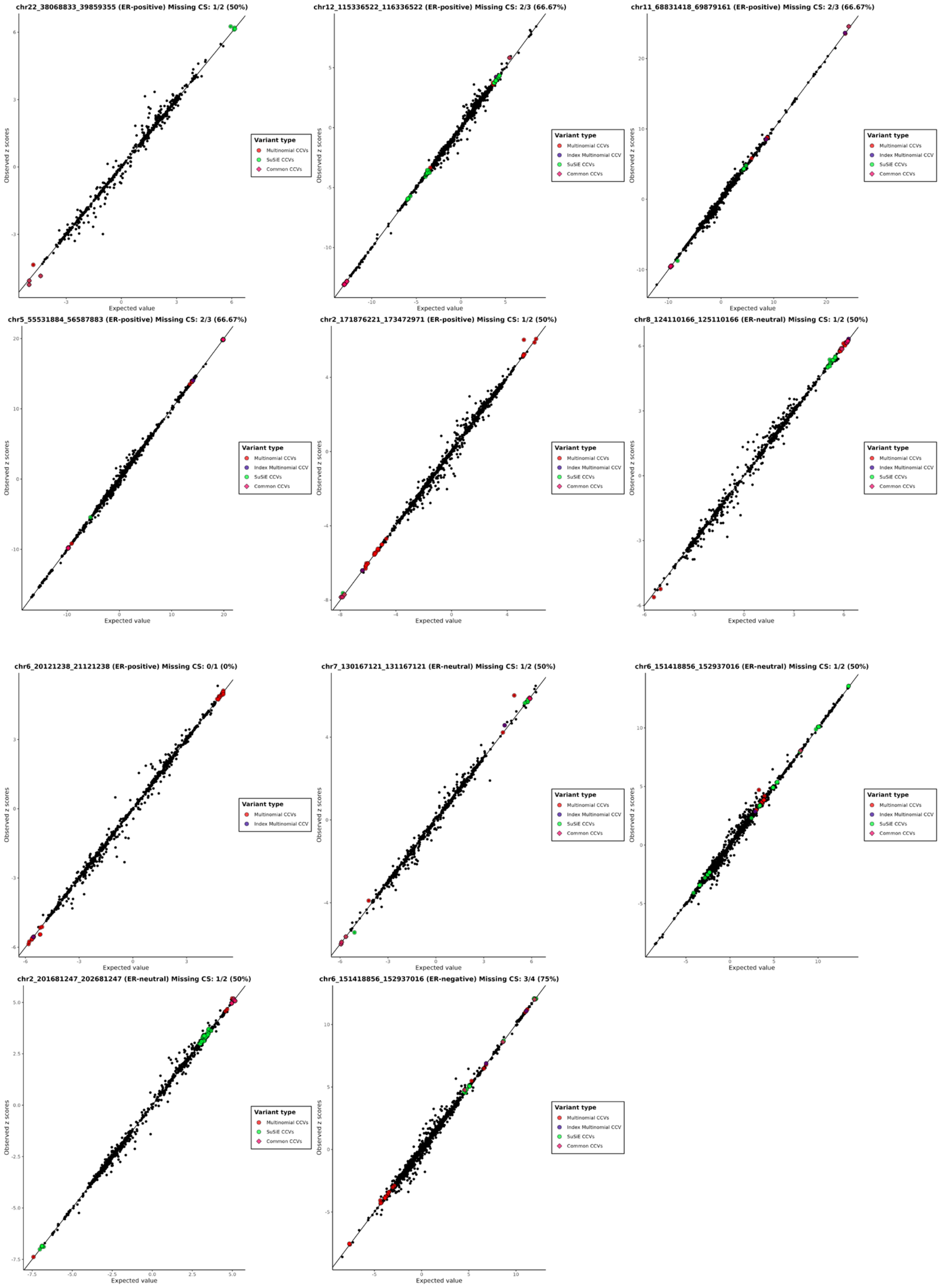


**Supplementary Figure 2. Expected vs observed z-score values for missing credible sets with *SuSiE* predictions.** Diagnostic plot created by the *SuSiE* function *fitted_rss*, assessing the correlation between the observed z-scores (summary statistics) vs expected (calculated from the LD matrix). Dots are colour coded: red for multinomial-nominated CCVs, purple for index multinomial CCVs, green for *SuSiE* CCVs, pink for overlapping CCVs between the methods. For each plot, the genomic region, ER-subtype for which the association was established and the credible set coverage is shown in the plot title.

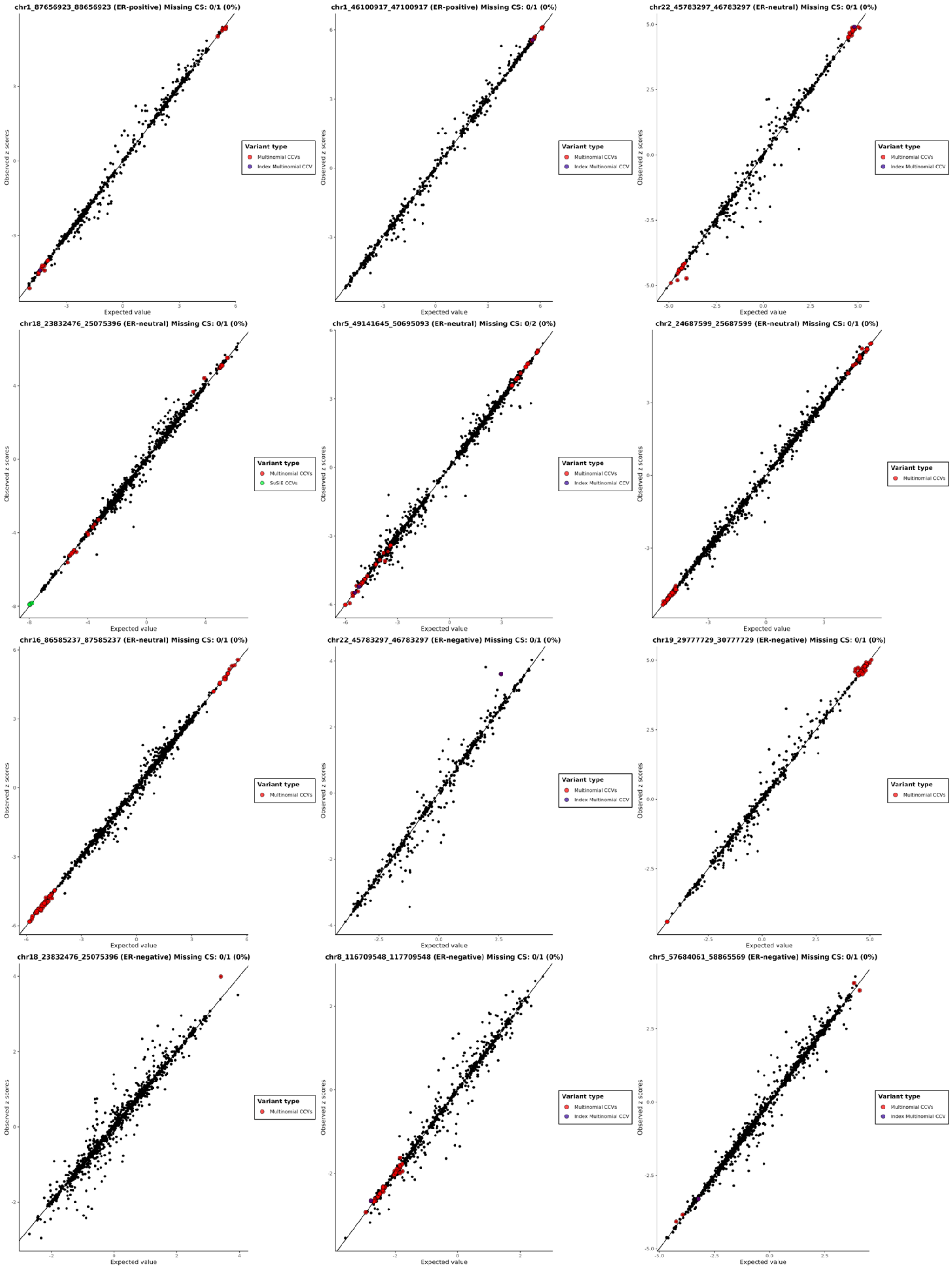


**Supplementary Figure 3. Expected vs observed z-score values for discordant credible sets with no *SuSiE* predictions.** Diagnostic plot created by the *SuSiE* function *fitted_rss*, assessing the correlation between the observed z-scores (summary statistics) vs expected (calculated from the LD matrix). Dots are colour coded: red for multinomial-nominated CCVs, purple for index multinomial CCVs, green for *SuSiE* CCVs, pink for concordant CCVs between the methods. For each plot the genomic region, ER-subtype for which the association was established and the % of credible sets found concordant between the methods are shown.

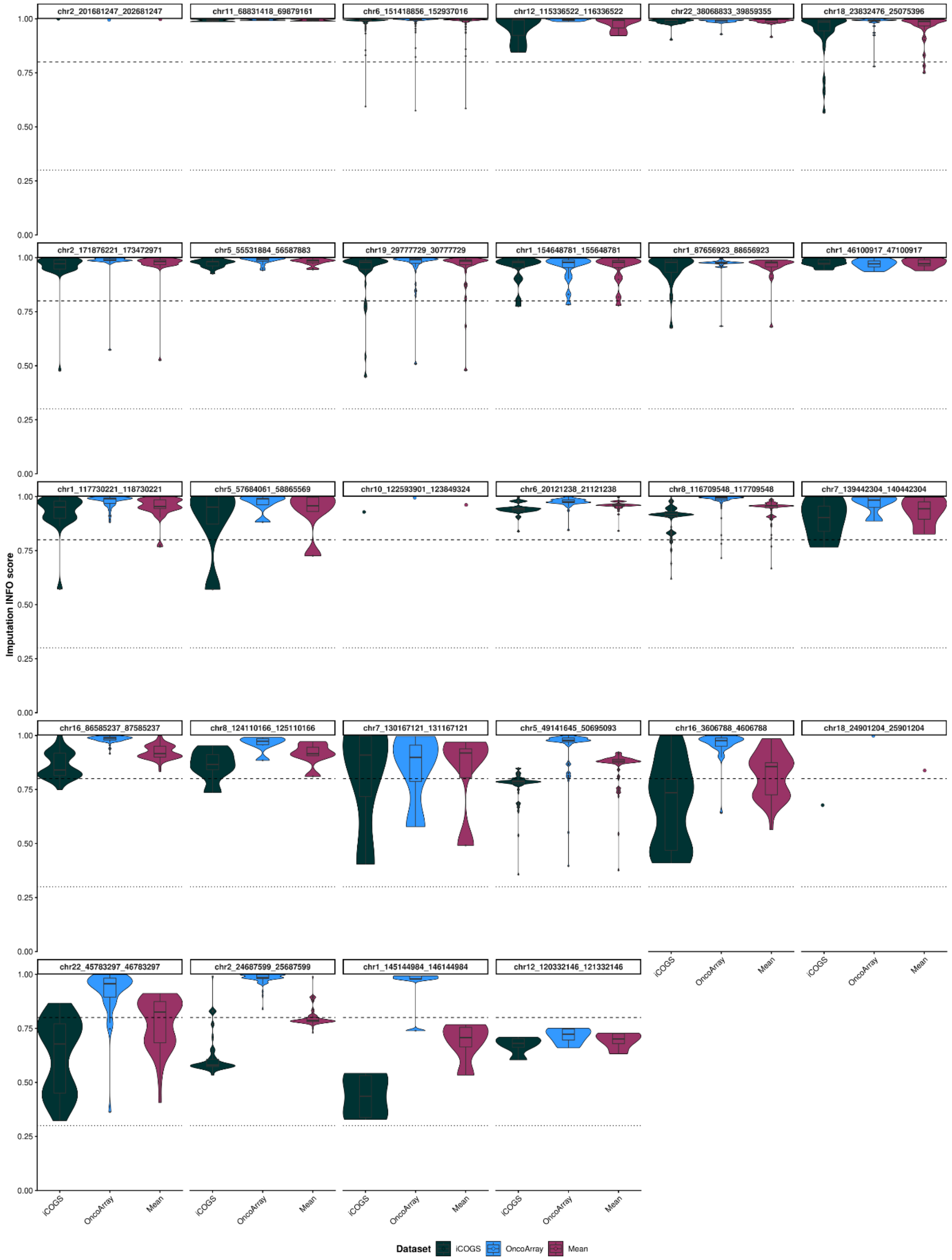


**Supplementary Figure 4. Imputation quality (INFO scores) for variants in missing credible sets.** Violin plots show the distribution of INFO scores across iCOGS, OncoArray, and their mean imputation quality for variants within regions where fine-mapping credible sets of Fachal *et al.* (2020) were not recovered. Each facet represents a genomic region, with datasets color-coded (iCOGS, dark teal; OncoArray, blue; Mean, deep pink). Horizontal dashed lines mark INFO = 0.8 (high-quality threshold) and INFO = 0.3 (low-quality threshold). Single points indicate regions with fewer than two variants.

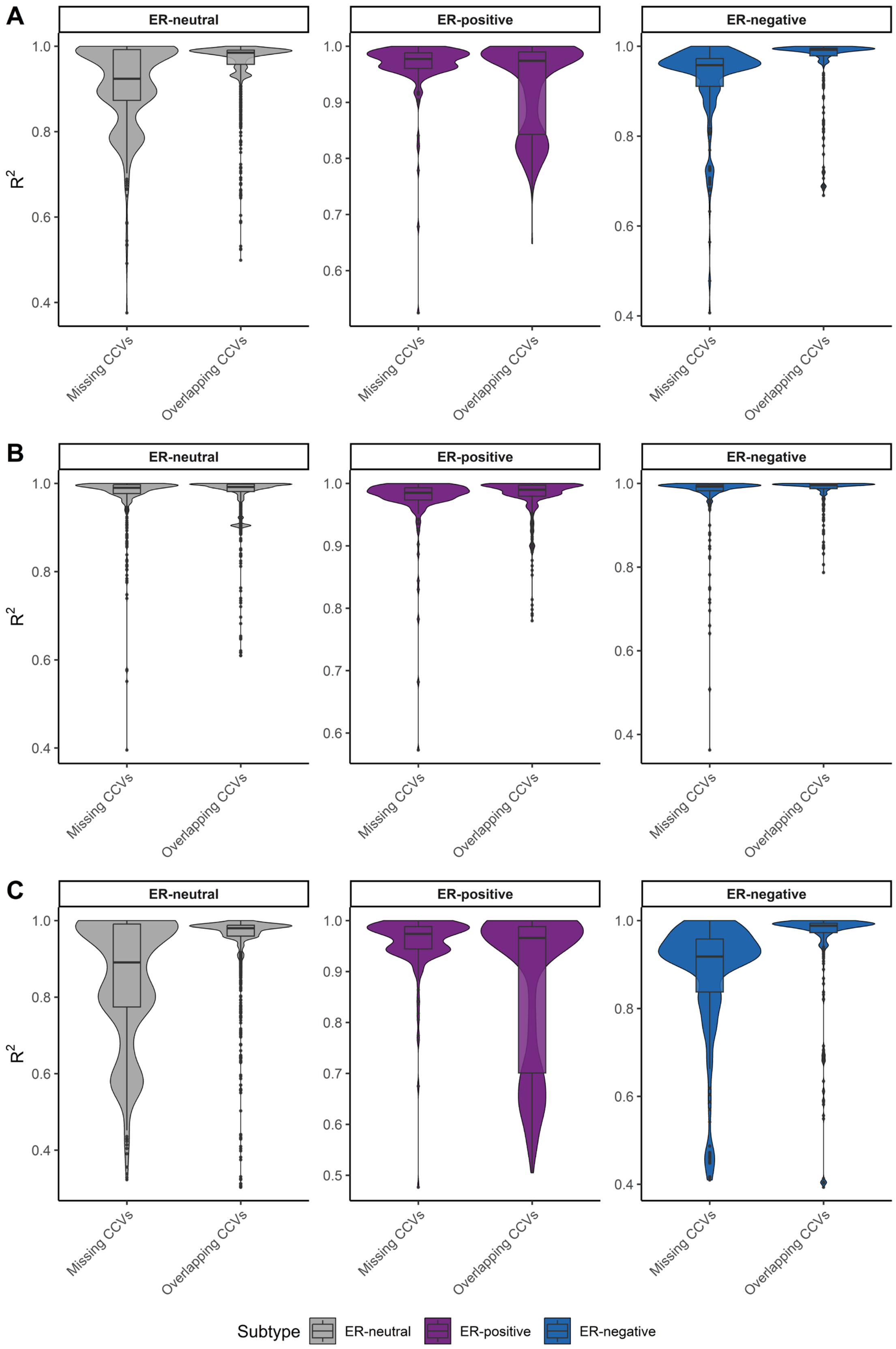


**Supplementary Figure 5. Imputation quality (INFO scores) for missing versus overlapping MNR-derived CCVs.** Violin plots show the distribution of INFO scores for (A) Mean info scores between iCOGS and OncoArray, (B) OncoArray info scores and (C) iCOGS info scores. The violin plots are colour-coded based on subtype.
